# Mining for Health: A Comparison of Word Embedding Methods for Analysis of EHRs Data

**DOI:** 10.1101/2022.03.05.22271961

**Authors:** Emily Getzen, Yucheng Ruan, Lyle Ungar, Qi Long

**Affiliations:** University of Pennsylvania, Philadelphia, PA 19104

## Abstract

Electronic health records (EHRs), routinely collected as part of healthcare delivery, offer great promise for advancing precision health. At the same time, they present significant analytical challenges. In EHRs, data for individual patients are collected at irregular time intervals and with varying frequencies; they include both structured and unstructured data. Advanced statistical and machine learning methods have been developed to tackle these challenges, for example, for predicting diagnoses earlier and more accurately. One powerful tool for extracting useful information from EHRs data is word embedding algorithms, which represent words as vectors of real numbers that capture the words’ semantic and syntactic similarities. Learning embeddings can be viewed as automated feature engineering, producing features that can be used for predictive modeling of medical events. Methods such as Word2Vec, BERT, FastText, ELMo, and GloVe have been developed for word embedding, but there has been little work on re-purposing these algorithms for the analysis of structured medical data. Our work seeks to fill this important gap. We extended word embedding methods to embed (structured) medical codes from a patient’s entire medical history, and used the resultant embeddings to build prediction models for diseases. We assessed the performance of multiple embedding methods in terms of predictive accuracy and computation time using the Medical Information Mart for Intensive Care (MIMIC) database. We found that using Word2Vec, Fast-Text, and GloVe algorithms yield comparable models, while more recent contextual embeddings provide marginal further improvement. Our results provide insights and guidance to practitioners regarding the use of word embedding methods for the analysis of EHR data.

## 1 Introduction

In 2009, the Health Information Technology for Economic and Clinical Health (HITECH) Act was passed as a means to promote the adoption and meaningful use of health information technology in United States [1]. As a result, the use of EHR systems has significantly increased in hospitals and other points of care over the past decade. Similar trends have also been observed in other countries. Currently, there are many diseases that go undiagnosed by a physician until they have already caused considerable damage to the body. Other diseases share common symptoms, making it difficult for physicians to differentiate them [2]. Flaws in clinical reasoning have also been shown to lead to diagnostic errors– potentially leading to harm for the patient and unnecessary medical expenses [3]. Going beyond human capability and harnessing the power of these rich datasets could lead to significant improvements in patients’ health and quality of life. EHRs that span years and detail the health history of patients are available for analysis, and the potential for their use in early stage disease detection, risk prediction, and treatment evaluation could prove to be an invaluable contribution to the advancement of precision health.

The original intention of recording medical data in EHR systems, however, was not for research purposes, but primarily for billing and data transfer, as indicated in [4]. As such, complex EHRs data oftentimes come from heterogeneous sources with characteristics that make it difficult to analyze with classical statistical and machine learning methods [2]. Some of these challenges include high dimensionality, the presence of structured and unstructured data, and time-series data collected at irregular intervals and frequencies (see Figure 1 for an example). Since data from individual patients are sequential in nature and have variable lengths, this makes it impossible to directly use classical dimension reduction and feature engineering methods such as principal component analysis [6] and factor analysis [7]. However, one way of extracting meaningful information from a complex sequential EHRs dataset is through the use of word embedding algorithms. In typical natural language processing settings, word embedding algorithms build a lower dimensional vector representation of a word from a corpus of text [5]. Two words that have a higher probability of appearing in the same sentence have a smaller cosine distance between their vector representations. These methods can be extended to EHRs by treating medical events or medical codes as “words”, and a collection of medical records as the “corpus”. The “context” around an event is defined as the order of medical events happening before and after that event [2]. Structured medical codes are an important piece of EHRs data and could contribute significantly to early-stage disease detection and risk prediction. Since they are sequential in nature, the features are not well-defined and dependencies exist between the events, thus making word embedding algorithms particularly effective tools for analysis.

**Fig. 1.**
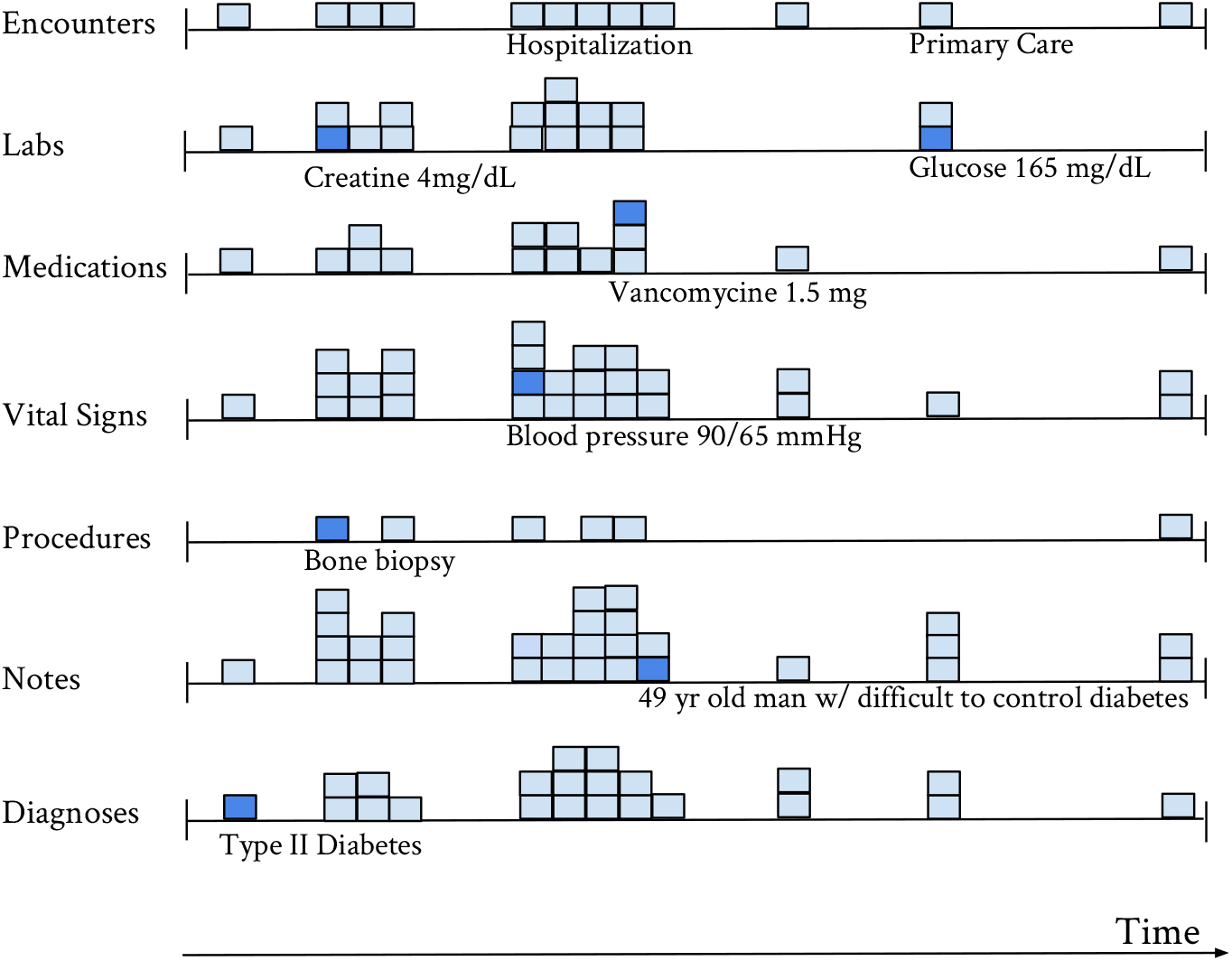
Sample patient health record data illustrating the challenges for analysis of EHRs data.

A growing body of literature explores the use of text mining and word embedding algorithms for analyzing EHR data. Miotto et al. (2016) sought to harness the rich information on patient data contained in clinical notes through deep learning, but raw text cannot be applied as meaningful features in such algorithms. After extracting words related to patient phenotype using standard ontologies for medical terminologies, the authors made use of word embedding algorithms like Word2Vec and Doc2Vec to convert the text into a form that the learning algorithms can understand. They also jointly embedded medical codes and words from clinical notes for EHR driven phenotyping [8]. Liu et al. (2015) utilized task-oriented resources to learn word embeddings for expanding ambiguous abbreviations in clinical notes, since the abbreviations are oftentimes difficult for both humans and machines to understand and utilize to improve patient care. In terms of accuracy, they achieved close to expert human performance [9]. Pham et al. (2016) embedded hospital admissions (diagnoses, procedures, and medications) into vectors, created a patient representation, and showed that the patient timeline vectors resulted in better predictive power than classifiers trained on raw data [10]. Word embedding can even be used to assist in de-identification of personal health information– Dernoncourt et al. (2016) created an automated de-identification system that generates embeddings for patient notes before feeding them into an artificial neural network (ANN) model [11]. Huang et al. (2019) developed ClinicalBERT which develops representations of clinical notes using bidirectional transformers [13], and Li et al. (2020) developed an embedding model known as BEHRT which employs 4 key concepts from an EHR: diseases, age, segment, and position [14].

The vast majority of work in this area, including the aforementioned papers, uses word-embedding methods for clinical notes in EHRs, but there has been very limited work on applying word embedding methods for the analysis of structured medical codes. Farhan et al. (2016) developed a sequential prediction model of clinical phenotypes by using word embedding vectors as features for their prediction model [2]. They used Word2Vec (skip-gram) to generate a vector representation of a structured medical code by feeding the algorithm with the medical code sequences. They then created vector representations of a patient’s entire medical history by multiplying each event’s vector by a temporal weight that was designed to account for the potential decay of impact of medical histories, and then summing across all the embeddings in a history. They proposed a prediction method called Patient-Diagnosis Projection Similarity (PDPS) in which they project patient vectors into the vector space and compute the cosine similarity between the patient vector and a diagnosis vector. The intuition behind the method is that the smaller the cosine distance, the more likely it is that the patient will get the diagnosis in the next visit. This allows for a multi-label classification problem. The authors found that PDPS achieved the highest performance in most cases when compared to logistic regression and other prediction methods. However, the logistic regression model did not incorporate the vector representations of medical events from Word2Vec [2]. Li et al. (2019) extended Word2Vec and PDPS to a distributed learning algorithm that can incorporate information from multiple data sources in one model without sharing individual-level data, and hence offer some level of data privacy protection [12].

One important limitation of [2] and [12] is that they used only Word2Vec to embed structured EHR events, but there is now a wide variety of word embedding tools such as FastText, GLoVe, ELMo, and BERT. In this work, we conduct a systematic comparison of re-purposing word embedding methods for analysis of structured medical data. By generating patient sequences using the same process as described in [2] and using the MIMIC-III database, we build multiple prediction models for the same diseases using different word embedding algorithms, and assess prediction performance and computation time. We also compare our disease-specific prediction models to the PDPS method in [2] to see if we can attain improved results. Our work seeks to compare the current state-of-the-art embedding methods and provide valuable insights on which types of methods are better.

## 2 Methods

We first briefly review popular word embedding methods, namely, Word2Vec [15], FastText [16], GloVe [17], ELMO [18], and BERT [19]. We then describe the approach for embedding EHRs data using the MIMIC III data as an example.

### 2.1 Word Embedding Methods

Word2Vec was proposed in 2013 by a research team led by Tomas Mikolov at Google. It is a two-layer neural network that is trained to convert words to vectors of real numbers such that the cosine distance between two vectors reflects the similarity of the two words. The input into the neural network is a corpus of text, and the output is a vector space with each unique word assigned a vector. It has two model formulations– continuous bag of words (CBOW) and skip-gram (see Figure 2). CBOW utilizes the surrounding context (other words around the target word) to predict the target word, and skip-gram uses a single word to predict a target context. The hidden layer following training contains the vectors corresponding to each word. Word2Vec depends on two parameters– size and context window. Size refers to the dimension of each output vector and context window determines the number of words before and after the target word to be included as context when training the algorithm. The original authors of Word2Vec recommend a context window of 10 for skip-gram and 5 for CBOW, and a size between 100 and 1,000.

**Fig. 2.**
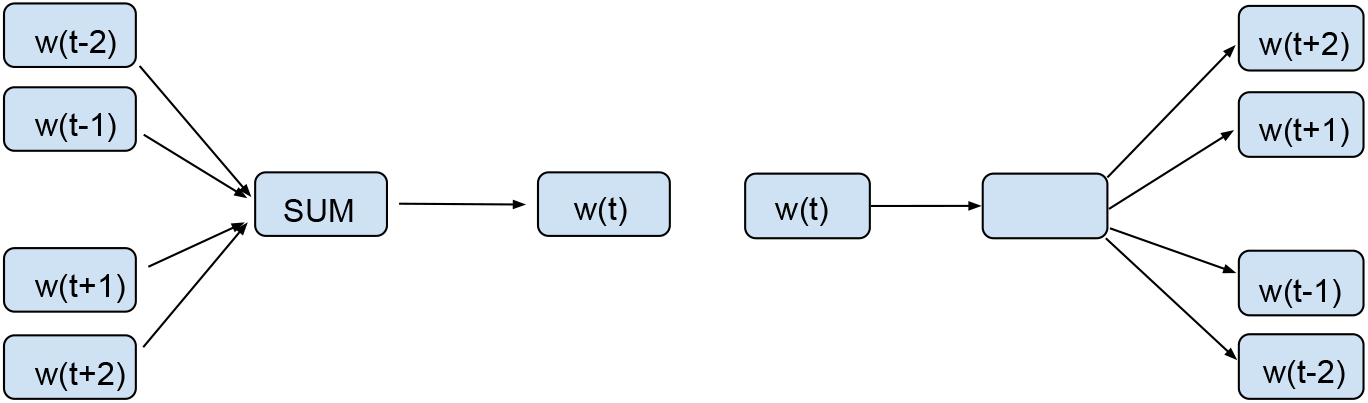
Two training mechanisms for Word2Vec and FastText: Continuous Bag of Words (CBOW) and Skip-Gram (w(t-i) refers to the ith word before the target word, w(t+i) refers to the ith word after the target word)

FastText is an extension of Word2Vec which also supports CBOW or skip-gram models. It was developed in 2015 by Facebook’s AI Research lab. The main difference is the fact that FastText represents each word as a collection of n-grams. The word “remember” for example, with n = 4, would have a FastText representation for the character n-grams as < *rem, reme, emem, memb, embe, mber, ber* >. The boundaries serve to protect the meanings of shorter words that might appear as character n-grams in a longer word, and are ignored during training. The algorithm generates a vector for each n-gram and sums across the vectors to get the resultant word embedding. The ability to more properly recognize rare words is a benefit of using character n-grams. With regard to medical data, it is unclear whether or not the same logic would hold since medical events are coded as individual entities without designated structures such as prefixes, suffixes, etc.

GloVe, or global vectors model, is an unsupervised learning algorithm developed in 2014 as an open-source project at Stanford. The main difference between GloVe and Word2Vec / FastText is that GloVe does not solely rely on local statistics (context windows used for training in CBOW and skip-gram) as it also incorporates global statistics (word co-occurence) to generate the vectors. GloVe derives the semantic relationships between words from a co-occurence matrix, which counts the number of times a given word has co-occured with another word. Table 1 provides an example on calculating the co-occurence matrix.

**Table 1.**
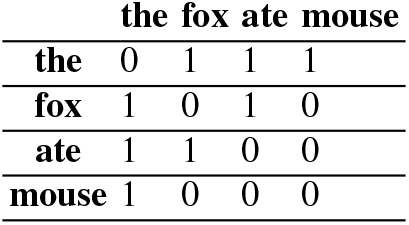
Sample co-occurence matrix for sentence “The fox ate the mouse” to illustrate GloVe training

The relationships between words is displayed by examining the ratio of cooccurence probabilities with probe words. For example, let *P*(*k* |*w*) be the conditional probability that word k appears in the context of word w. The word “ice” tends to co-occur more frequently with the word “solid” as opposed to “gas”, whereas the word “steam” tends to co-occur more frequently with “gas” rather than “solid”, but both words co-occur frequently with the word “water” and infrequently with the word “fashion”. So for k = solid, *P*(*k* |*ice*) will be high, *P*(*k* |*steam*) will be low, and the ratio between the two will be very large. For k = water or k = fashion, the ratio will be close to 1; and for k = gas, the ratio will be very small [17]. The training objective is to learn word vectors such that the dot product equals the logarithm of words’ probability of co-occurence. The logarithm of the ratios is associated with the vector differences in the word vector space, thus capturing more information. Table 2 provides an example on calculating log ratios of word co-occurences.

**Table 2.** Log ratios of word co-occurences from a sample corpus of text to illustrate how distances between word vectors in space are determined.

| Probability and Ratio | k = solid | k = gas | k = water | k = fashion |
| --- | --- | --- | --- | --- |
| $P(k \mid \text{ice})$ | $1.9 \times 10^{-4}$ | $6.6 \times 10^{-5}$ | $3.0 \times 10^{-3}$ | $1.7 \times 10^{-5}$ |
| $P(k \mid \text{steam})$ | $2.2 \times 10^{-5}$ | $7.8 \times 10^{-4}$ | $2.2 \times 10^{-3}$ | $1.8 \times 10^{-5}$ |
| $P(k \mid \text{ice}) / P(k \mid \text{steam})$ | 8.9 | $8.5 \times 10^{-2}$ | 1.36 | 0.96 |

ELMo word embeddings are learned from a deep bidirectional language model (biLM) that is pre-trained on a large text corpus. The mechanism seeks to address challenges in both modeling complex characteristics of words (syntax and semantics) as well as linguistic contexts (modeling polysemy). Unlike the other methods listed above, ELMo representations are deep– they are a function of the internal layers of the biLM (see Figure 3). The higher level LSTM architecture captures more context-dependent parts of word meanings and the lower levels capture more of the aspects of syntax. It is a character-based model which further allows it to form representations of out-of-vocabulary words. ELMo can be customized for different NLP tasks by taking a linear combination of all hidden states of LSTM, with weights being task-specific (for example, one linear combination for a next word prediction task, and another linear combination for a text summarization task).

**Fig. 3.**
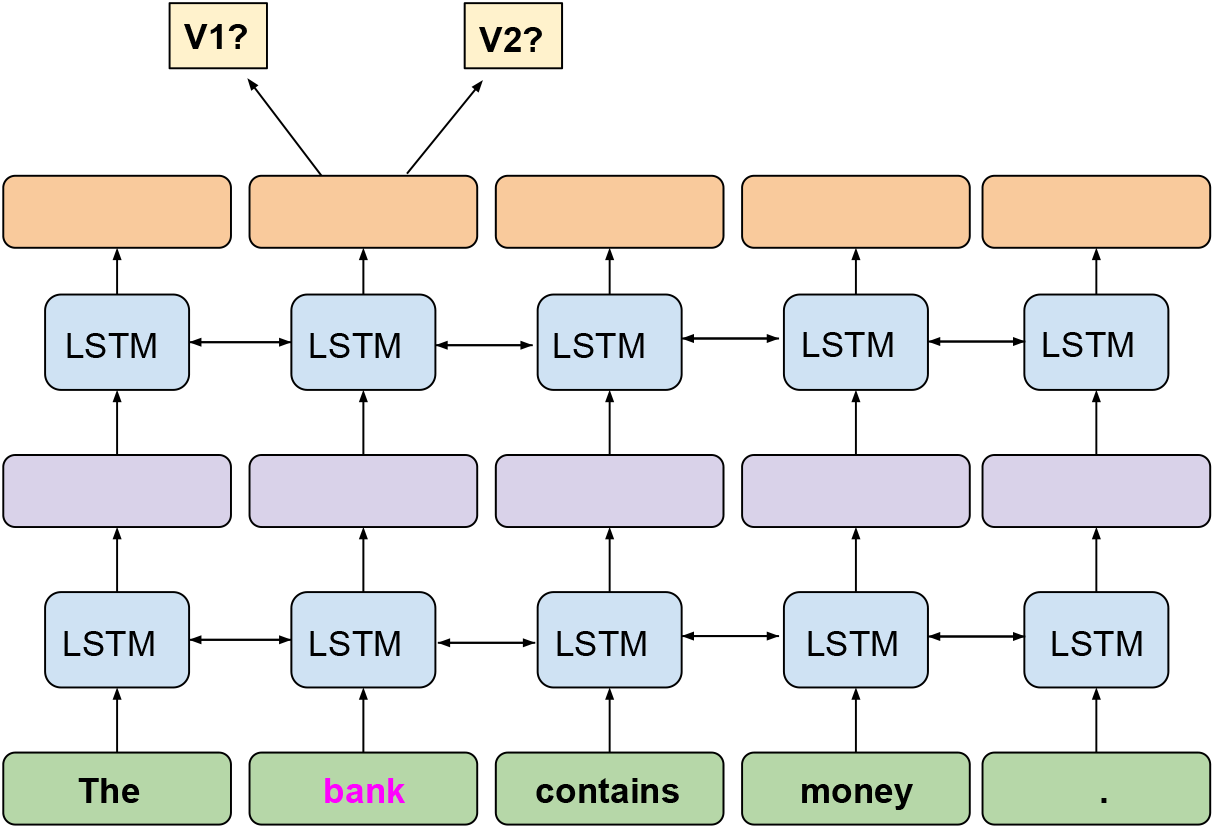
ELMo bidirectional LSTM training mechanism can produce different embeddings for the same word depending on the context

BERT, or Bidirectional Encoder Representations from Transformers, is a word embedding algorithm developed by Google in 2018. It incorporates a transformer neural network architecture– an encoder-decoder model which utilizes attention mechanisms. This architecture has been shown to have advantages over other sequential models such as LSTM, RNN, etc. BERT uses masked language modeling for training, meaning it masks 15% of the words in a sequence and uses the position information to infer what they should be (see Figure 4). The BERT model uses 12 layers of transformer encoders and each output can be used as a word embedding (the authors identified that the best approach was to sum the last four layers). Out of the five embedding methods described, BERT is the only algorithm that can read the sentence bidirectionally since its self attention layer performs self attention on both directions.

**Fig. 4.**
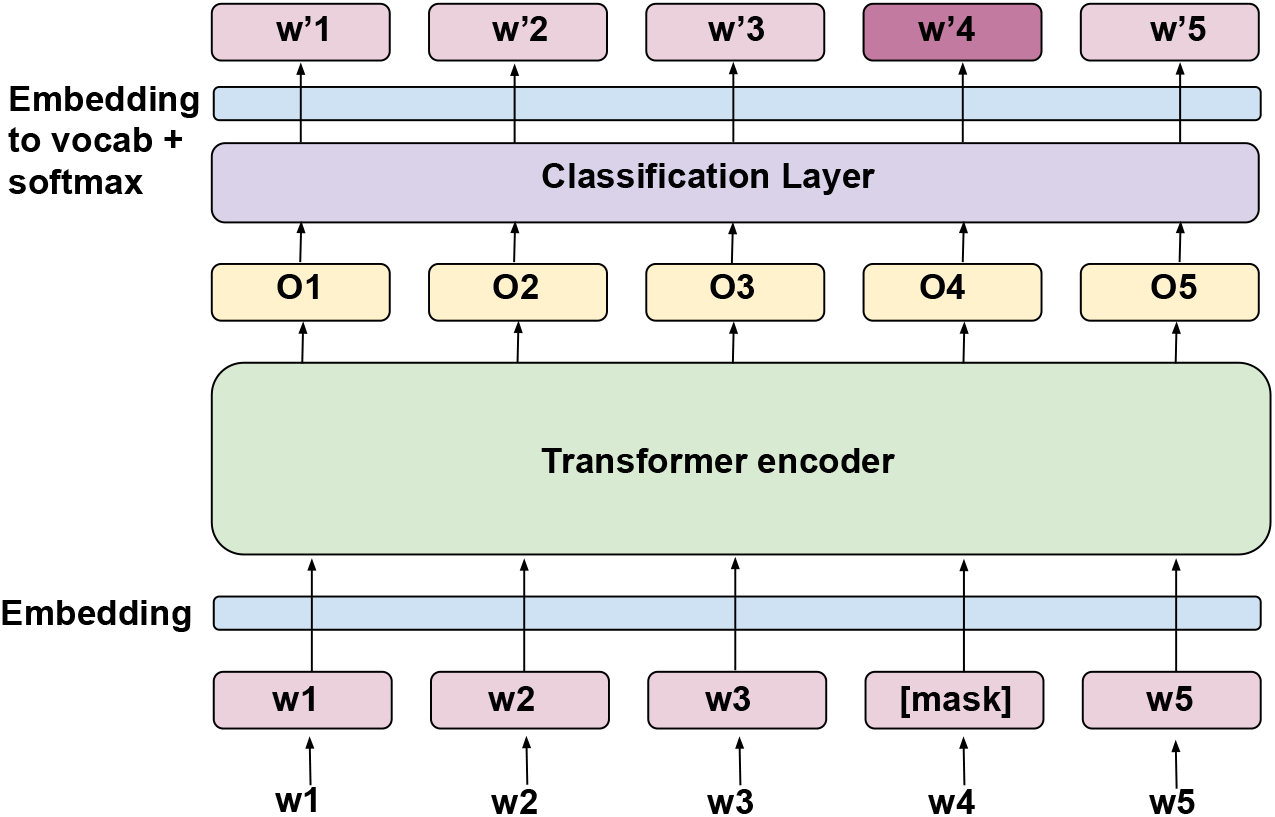
To train, BERT masks a percentage of words and tries to predict them

There are two types of word embeddings that can be generated using a word embedding algorithm– static word embeddings and contextual word embeddings. Models that generate static embeddings create a single representation for a word. This is oftentimes problematic in languages since polysemous words must share a single vector. Word2Vec, FastText, and GloVe can only create static embeddings. Contextual embeddings, in contrast, are also a function of the “context” of the word–e.g., the other words in the sentence. This allows the vector representation for a word to differ depending on its meaning, as inferred from its context. For example, for the two sentences “let’s swim by the river bank” and “I have to deposit money at the bank”, the embedding for the word “bank” would differ for the two sentences since it has different meanings based on the context. Both ELMo and BERT have the ability to generate context-dependent word embeddings, on top of static embeddings. Contextual embeddings could prove to be useful with regard to medical data, since depending on how the physician input the codes, some ICD-9 codes could be generalized to levels which can lack some disease specificity (level 3 diagnoses could represent types of diseases and level 5 diagnoses could represent specific diseases). An example is hyperthyroidism and hypothyroidism falling under the same category when generalized to level 3. Contextual embeddings could help distinguish these diagnoses based on the medical context.

### 2.2 Embedding of EHRs

We used the Medical Information Mart for Intensive Care III (MIMIC III) database from [20], which consists of de-identified health-related data of patients who stayed in critical care units. There were 3950 patients in the training set and 1692 patients in the test set. For each medical record we extracted codes for prescriptions, lab tests, symptoms, conditions, and diagnoses. The mean number of structured medical codes per patient was 315, with a minimum number of events of 7, and a maximum of 3463 (see Figure 5 for distribution of events). The maximum number of medical visits in a patient’s record is 42 (see Figure 6 for distribution of medical visits). In using this data for word embedding, one can think of the “word” as a structured medical code in the patient record (a prescription, lab test, symptom, condition, or diagnosis), and a “sentence” as the chronological sequence of all structured events in a patient record (see Figure 7 for data structure). Since we wish to predict future diagnoses, we split the sequence of events into the feature window and prediction window. We define target diagnosis as the ICD9 diagnosis code corresponding to the disease that we want to predict. If the target diagnosis is present in a patient’s medical record, the feature window contains the sequence of structured medical events up to the visit that contains the first target diagnosis. If the target diagnosis is not present in a patient’s medical record, the feature window contains all medical visits up to the most recent visit. The prediction window comprises either the most recent medical visit, or the visit containing the first occurrence of the target diagnosis. It is positive if it contains the target diagnosis, and negative if it does not contain the target diagnosis. The events in the feature window are used to generate the vector representation of the patient history (see Figure 8 for the workflow).

**Fig. 5.**
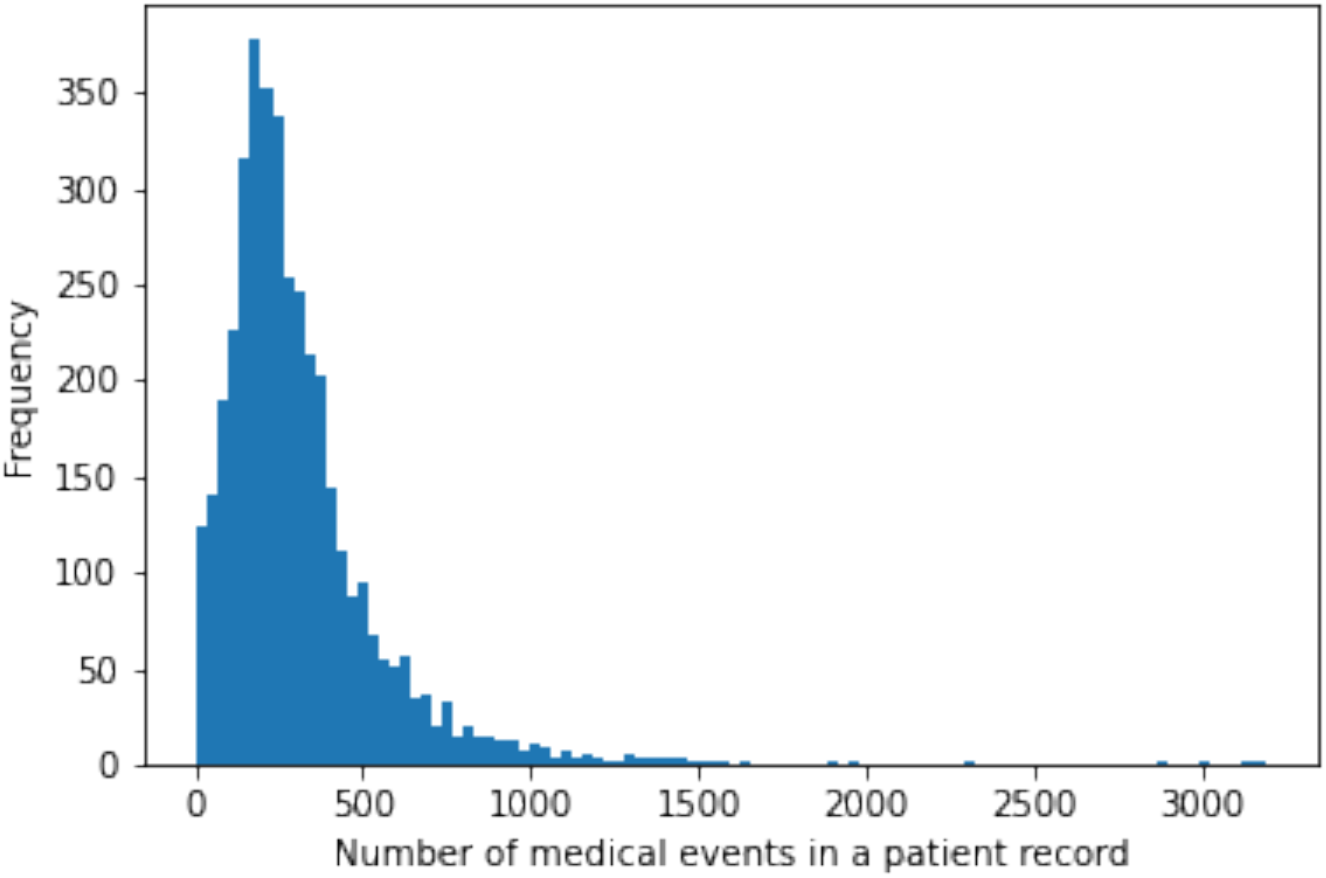
Distribution of number of structured medical codes per patient medical record.

**Fig. 6.**
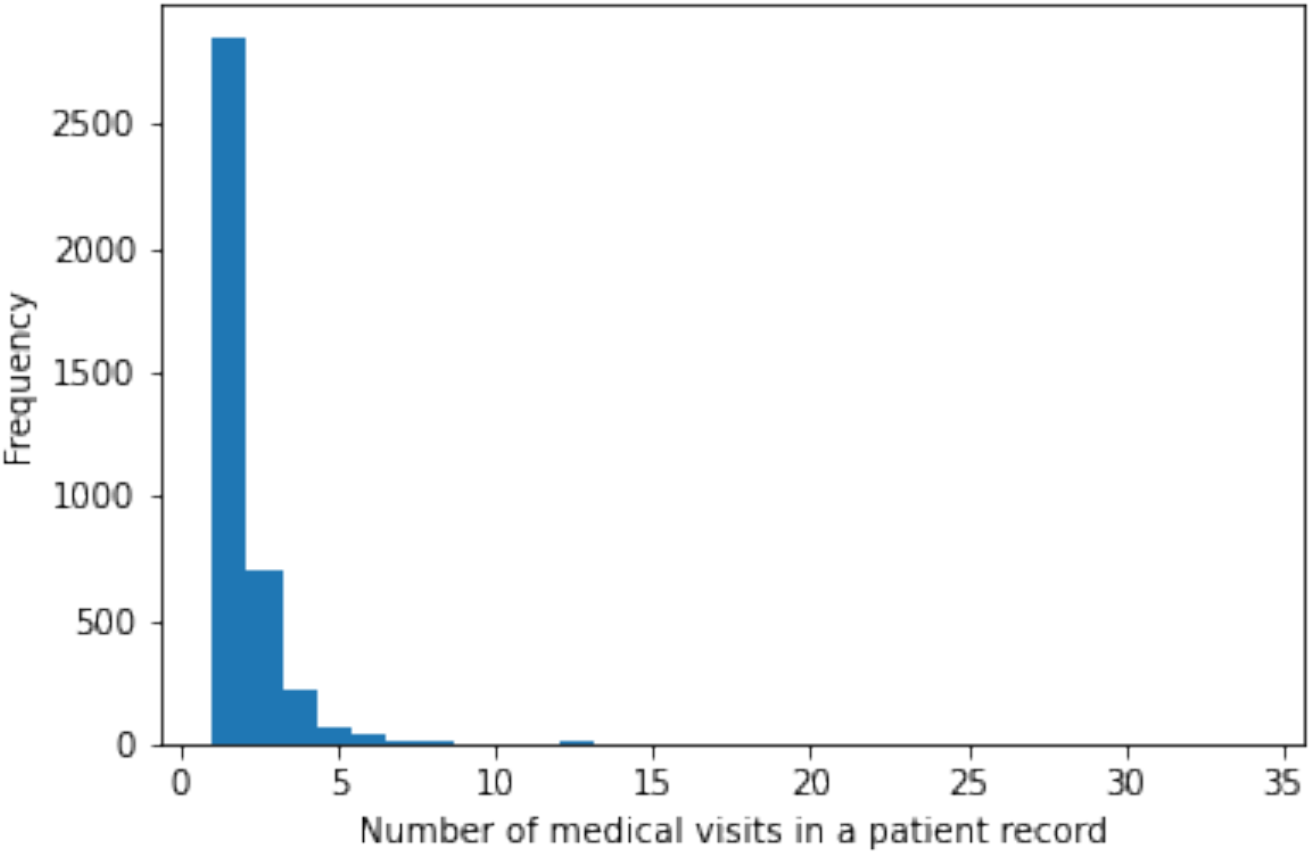
Distribution of number of medical visits per patient medical record.

**Fig. 7.**
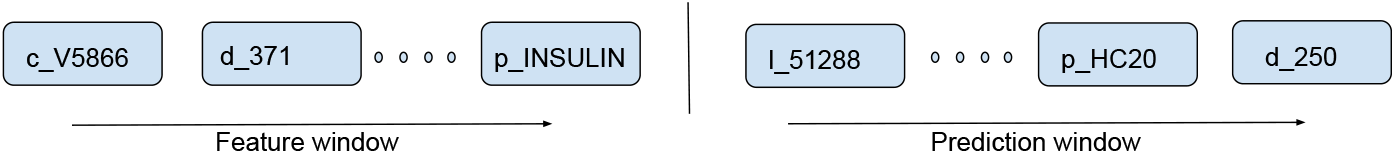
MIMIC-III structured event data structure example. The structured events in an individual’s medical record are split into the feature window and prediction window.

**Fig. 8.**
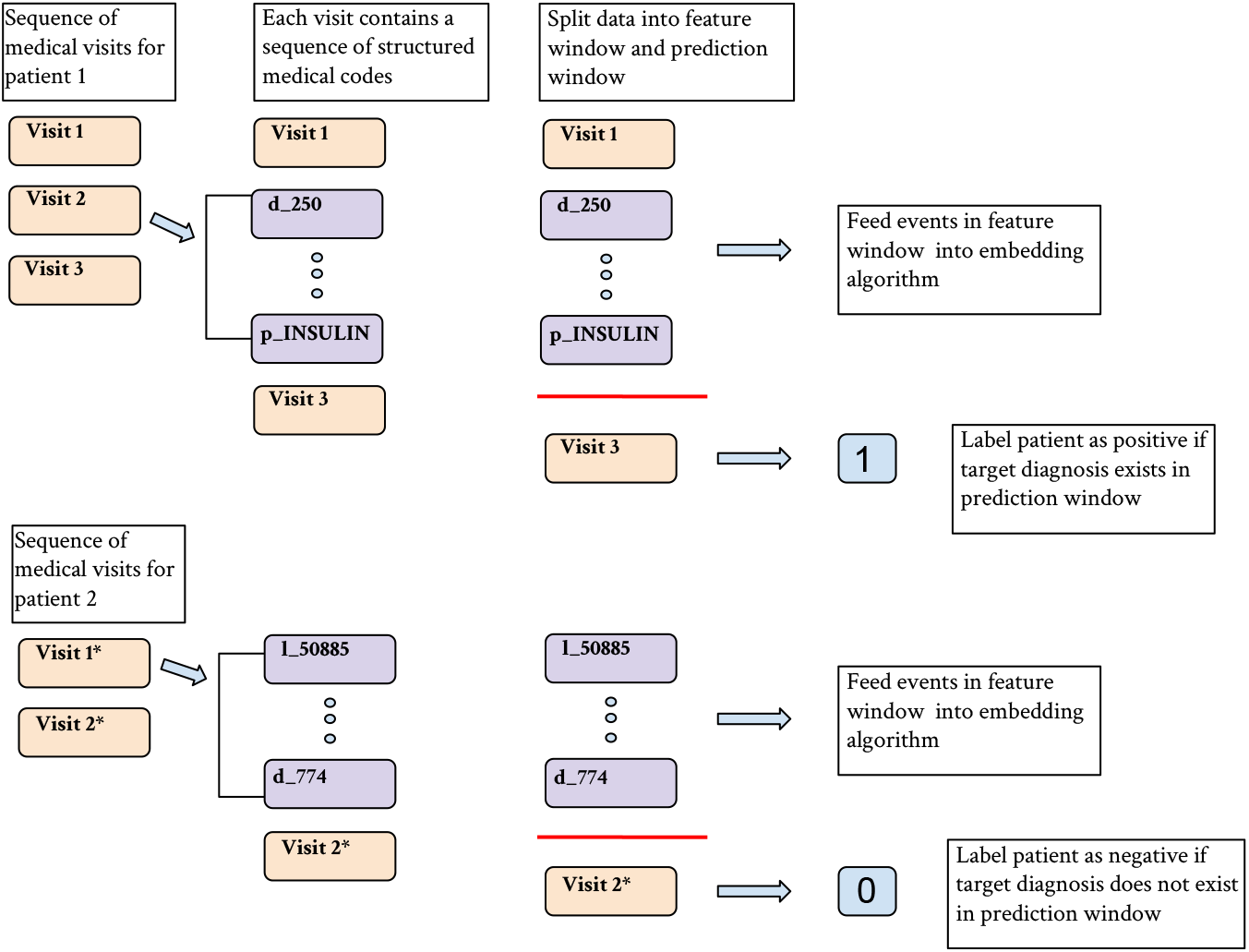
Each patient may have a different number of medical visits that occur at different times (* indicates different visit time). Each medical visit has a variable sequence of medical codes indicating prescriptions, lab tests, diagnoses, conditions, or symptoms. Medical events prior to the visit containing the diagnosis of interest or the most recent visit comprise the feature window. These events are fed into the word embedding algorithm. The remaining visit is the prediction window and determines whether or not a patient is labeled positive or negative for having the diagnosis of interest present in this visit.

There are a few ways to create an overall vector representation of the patient history. We follow [2] which gives more recent events greater weights in the overall representation by multiplying each event by a temporal weight. They then sum the weighted embeddings for each event to obtain the overall vector representation of the patient history to be used as the features for prediction (see Figure 9). The equation that describes this process is as follows:

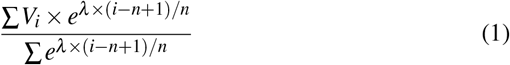

where *V*_*i*_ is the embedding for event *i* in a patient’s medical record, *λ* is the time decay factor, and *n* is the total number of events in the record. For our experiments, a decay factor of 5 is used.

**Fig. 9.**
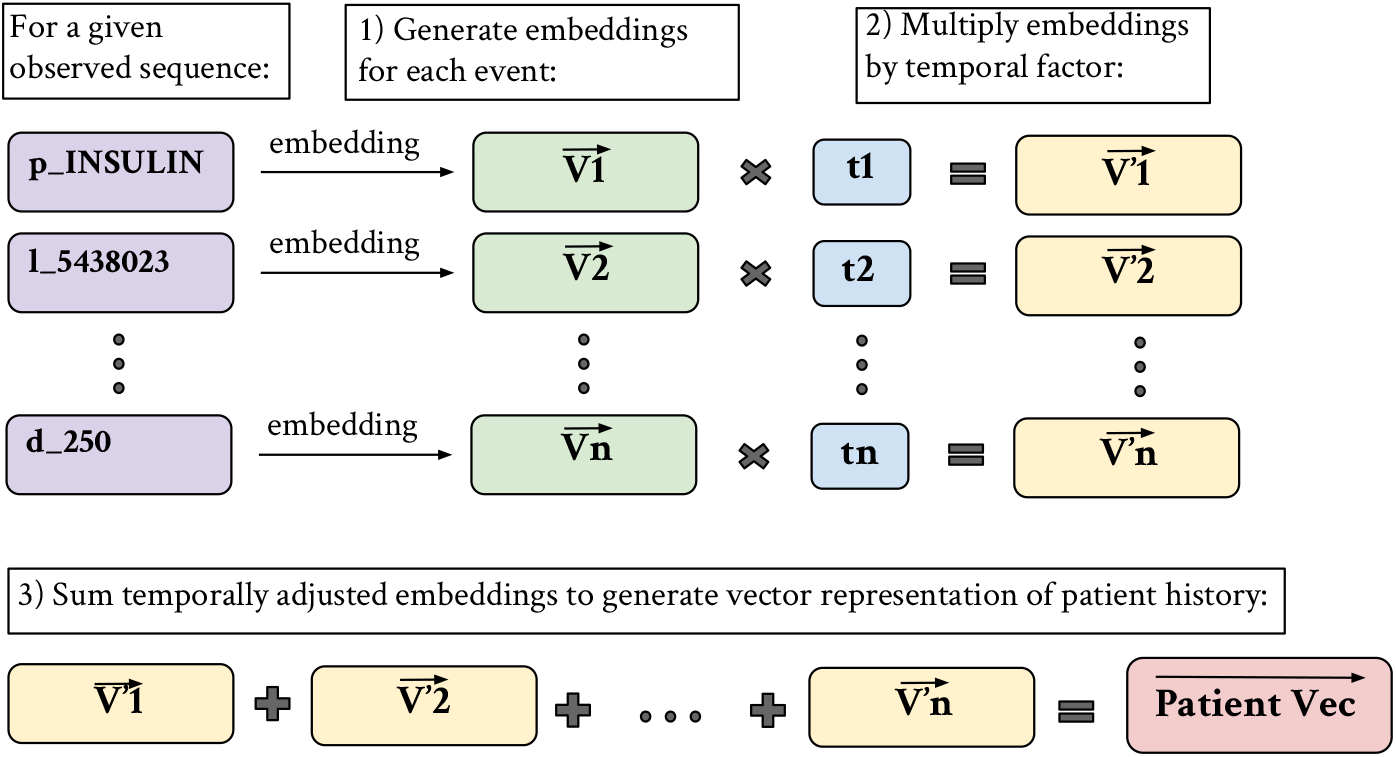
Patient vector generation via temporal summation of individual event embeddings. 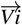 refers to the embedding for event i, ti refers to the temporal weight applied to its corresponding event Embedding, 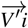 refers to the resultant embedding following temporal adjustment, 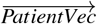 refers to the vector sum of the adjusted events in a patient history

Overall patient representations can also be generated without the temporal weights. This involves simply obtaining the embeddings for each medical event and summing across all of the embeddings in the patient record.

## 3 Numerical Experiments

To evaluate the performance of the embedding algorithms described in Section 2, we use the embeddings of patient medical history to build prediction models for disease diagnoses using EHRs data prior to the time of diagnosis and compare prediction performance.

### 3.1 Building Disease Prediction Models

To build disease prediction models, we first split patient medical records into the patient history (events prior to the most recent visit) and the events in the most recent visit (see Figure 7). The outcome for the patient is labeled as positive if the target diagnosis is present in the most recent visit. If the target diagnosis is present in the previous medical history, we only use the chronological sequence of events leading up to the visit containing the first diagnosis, and exclude the subsequent visits to prevent the target itself being used as a feature in prediction for the next visit.

Farhan et al. (2016) developed a prediction model known as Patient Diagnosis Projection Similarity (PDPS) (see Figure 10) [2]. This prediction model involves projecting patient sequences into the vector space while accounting for the temporal impact of events as described in the previous section. The method uses the cosine similarity between the projected patient vector and the diagnosis vector to predict whether or not the patient is at risk for developing that disease. The main benefit that this method provides is its ability to predict risks for multiple diseases simultaneously.

**Fig. 10.**
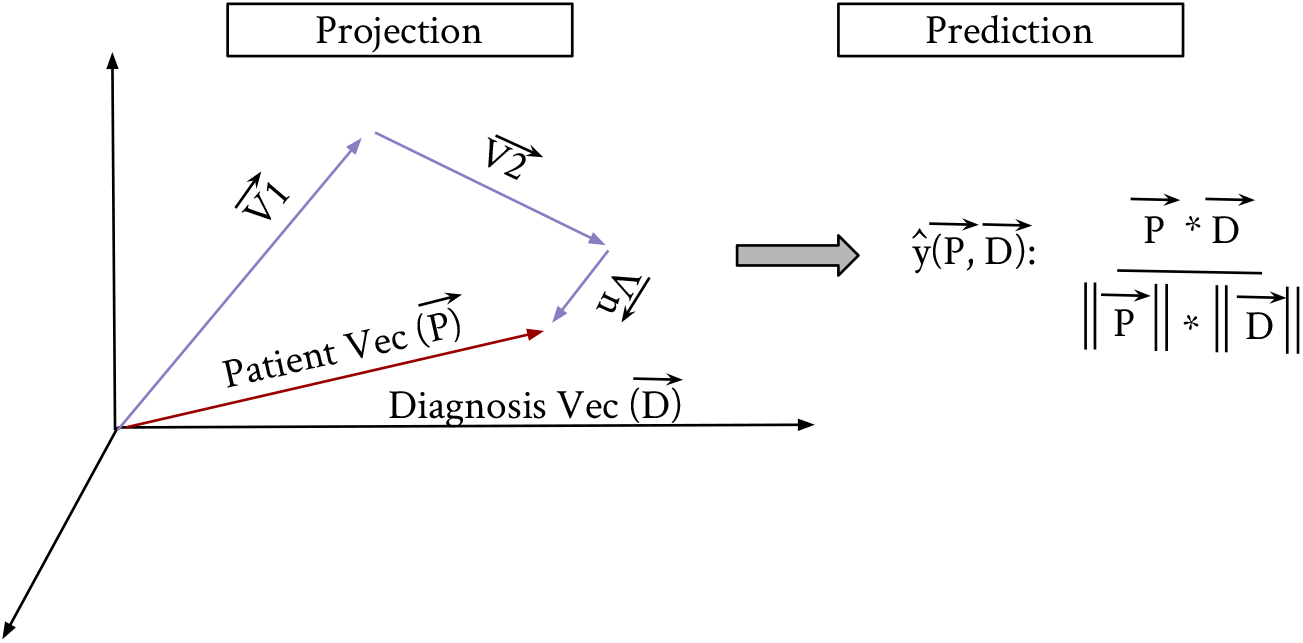
Patient Diagnosis Projection Similarity (PDPS) involves predicting a future diagnosis based on the angle between the patient vector and the diagnosis vector. 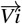 refers to the embedding for event i, 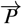 refers to the patient vector following weighted event vector summation, 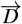 refers to the vector for the target diagnosis that we want to predict in the next visit, and 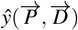 refers to the cosine similarity of the patient vector and target diagnosis vector

However, improvement in prediction accuracy could be achieved with building disease-specific prediction models instead. We develop such models based on each aforementioned word embedding algorithm using both penalized logistic regression and deep learning methods. Briefly, we use each embedding algorithm to generate a patient history vector representation of dimension 100, where each element of a patient vector is used as a feature in prediction. We assess embedding algorithms by predicting a variety of diagnoses, and comparing the model performance.

We assess model performance using both F1 score and AUC. The equation for F1 score is as follows :

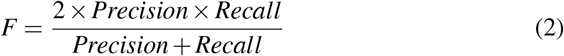

where

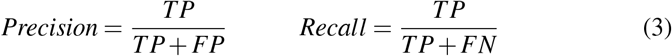

Our experiments focus on predicting diseases that have a prevalence of > 0.08 in the MIMIC dataset. We include 6 of those diagnoses with varying prevalence in our results (see Table 3). These diagnoses are: Diabetes, Chronic Kidney Disorder, Heart Failure, Lipid Metabolism Disorder, Fluid / Electrolyte Disorder, and Cardiac Dysrhythmia.

**Table 3.**
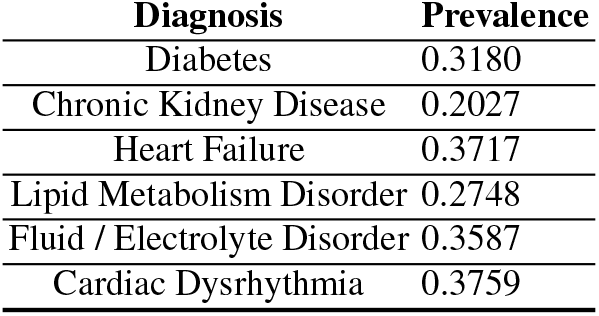
Diagnoses whose presence in the next visit is used as outcome variable in the numerical experiments, and their prevalence in the data

### 3.2 Experimental Set Up

All numerical experiments are carried out using Python version 3.6 and Tensorflow version > 1.10. To obtain the processed MIMIC-III sequences containing only diagnoses, prescriptions, lab tests, symptoms, and conditions, we first follow the instructions to install MIMIC-III in a local Postgres database using the tutorial on the MIMIC website. We then follow the data pre-processing steps from the SequentialPhenotypePredictor github page from [2]. We use the Gensim package to train Word2Vec and FastText, and the package glove python from pypi to train our GloVe model. We train our medical ELMo model utilizing a Tensorflow implementation using the bilm-tf github page from [18]. As for the medical BERT pre-training process, we follow the instructions on the official BERT github [19]. We first transform the plain text corpus of structured medical data into a serialized format suitable for training in tensorflow. Using the provided scripts, we learn the WordPiece vocabulary and pre-train BERT models on our GPU and CPU servers separately. To extract the embeddings from the pre-trained BERT models, we can use the Python packages, bert-serving-server and bert-serving-client, to obtain the sentence (word) encodes from our pre-trained models. After completing embedding using one of the above-mentioned algorithms, we use the sklearn Python module to build our lasso prediction model (referred to as “Lasso” in our tables), and tensorflow.keras to build our artificial neural network prediction model with two hidden layers (referred to as “DL” in our tables). We do 50 runs for each method and average each AUC and F1 score. These averages are presented in the subsequent tables.

### 3.3 Hyperparameters

Our Word2Vec model was trained with the skip-gram method with a context window of 10, and a size of 100, and 70 epochs. Our FastText architecture uses the same parameters as the Word2Vec model. Our GloVe architecture trains with a window size of 10, 100 components, and 70 epochs. Our ELMo architecture trains using the bidirectional mechanism with 7 filters for the character CNN portion, 10 percent dropout, 2 LSTM layers with dimension 1024 and projection dimension 50, 10 epochs, and batch size of 128. Our BERT architecture has 12 hidden layers, 10 attention heads, 10 percent dropout and yields an output embedding dimension of 100. Our feed forward neural network (”DL”) used for evaluation has two hidden layers. The first layer has 55 nodes and the second has 20. It uses mean squared error loss function and adam optimizer.

### 3.4 Results

We first compare model performance from the word embedding algorithms using static embeddings only. Recall, Word2Vec, FastText, and GloVe can only produce static embeddings, whereas ELMo and BERT can produce both static and contextual embeddings. For ELMo and BERT, we generate static embeddings for each unique medical event in the data by treating each event as its own sentence so as not to incorporate contextual information. In our first set of results, we incorporate the temporal decay into the construction of patient sequences (see Table 4 and Figure 12, Figure 14, and Figure 16).

**Table 4.** Prediction performance for various models that use patient vectors generated by each embedding algorithm based on static embeddings and temporal adjustment

| **Diabetes** | **Word2Vec** | **FastText** | **GloVe** | **ELMo** | **BERT** |
| --- | --- | --- | --- | --- | --- |
| AUC (lasso) | 0.7588 | 0.7451 | 0.7467 | 0.7243 | 0.6624 |
| AUC (DL) | 0.7444 | 0.7547 | 0.7329 | 0.6665 | 0.6028 |
| F1 score (lasso) | 0.0825 | 0.0843 | 0.1020 | 0.0810 | 0.0204 |
| F1 (DL) | 0.1244 | 0.1499 | 0.1651 | 0.1542 | 0.0088 |
| **Chronic Kidney Disease** |  |  |  |  |  |
| AUC (lasso) | 0.7596 | 0.7479 | 0.7328 | 0.7234 | 0.6772 |
| AUC (DL) | 0.7951 | 0.7797 | 0.7859 | 0.7079 | 0.6620 |
| F1 score (lasso) | 0.1067 | 0.0946 | 0.0403 | 0.0284 | 0.0007 |
| F1 score (DL) | 0.1231 | 0.1161 | 0.1726 | 0.1089 | 0.0000 |
| **Heart Failure** |  |  |  |  |  |
| AUC (lasso) | 0.7201 | 0.7283 | 0.7282 | 0.6792 | 0.6739 |
| AUC (DL) | 0.7313 | 0.7329 | 0.7033 | 0.6594 | 0.6522 |
| F1 score (lasso) | 0.1757 | 0.1993 | 0.2215 | 0.1404 | 0.1478 |
| F1 score (DL) | 0.2770 | 0.2793 | 0.2951 | 0.2295 | 0.0116 |
| **Lipid Metabolism Disorder** |  |  |  |  |  |
| AUC (lasso) | 0.7211 | 0.6950 | 0.7013 | 0.6600 | 0.6503 |
| AUC (DL) | 0.7072 | 0.7107 | 0.7038 | 0.6669 | 0.6447 |
| F1 score (lasso) | 0.1277 | 0.0885 | 0.0873 | 0.1086 | 0.0442 |
| F1 score (DL) | 0.1097 | 0.0761 | 0.1799 | 0.1647 | 0.0157 |
| **Fluid / Electrolyte Disorder** |  |  |  |  |  |
| AUC (lasso) | 0.6510 | 0.6317 | 0.6507 | 0.6194 | 0.5909 |
| AUC (DL) | 0.6361 | 0.6474 | 0.6349 | 0.5976 | 0.6003 |
| F1 score (lasso) | 0.3701 | 0.3723 | 0.3813 | 0.3093 | 0.3083 |
| F1 score (DL) | 0.4201 | 0.4105 | 0.4149 | 0.4075 | 0.1182 |
| **Cardiac Dysrhythmia** |  |  |  |  |  |
| AUC (lasso) | 0.6317 | 0.6353 | 0.6360 | 0.6175 | 0.6077 |
| AUC (DL) | 0.6400 | 0.6566 | 0.6152 | 0.5887 | 0.5758 |
| F1 score (lasso) | 0.0502 | 0.0641 | 0.0900 | 0.0568 | 0.0145 |
| F1 score (DL) | 0.1090 | 0.0818 | 0.1295 | 0.1950 | 0.0011 |

**Fig. 11.**
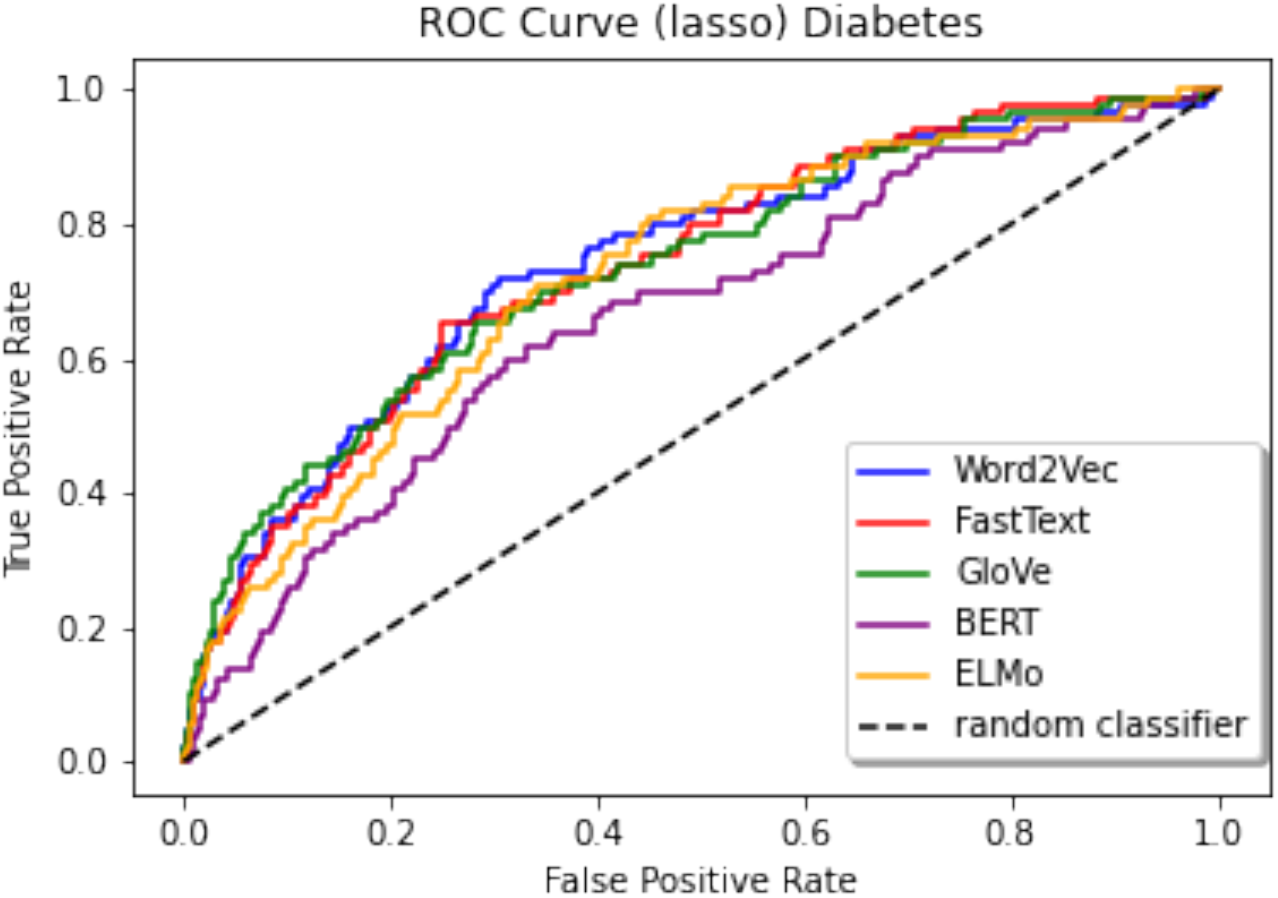
ROC curves for predicting Diabetes (lasso)

**Fig. 12.**
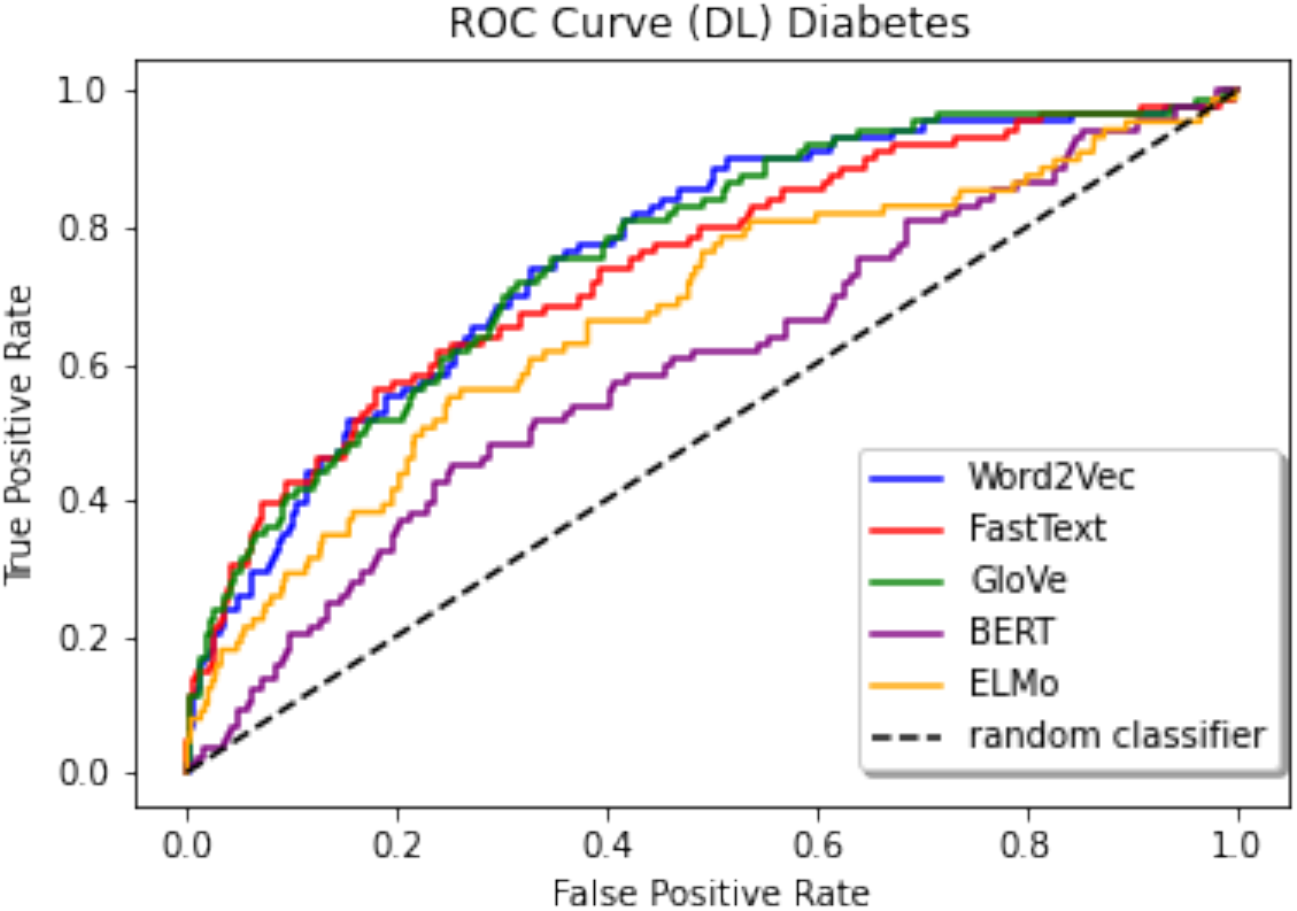
ROC curves for predicting Diabetes (DL)

**Fig. 13.**
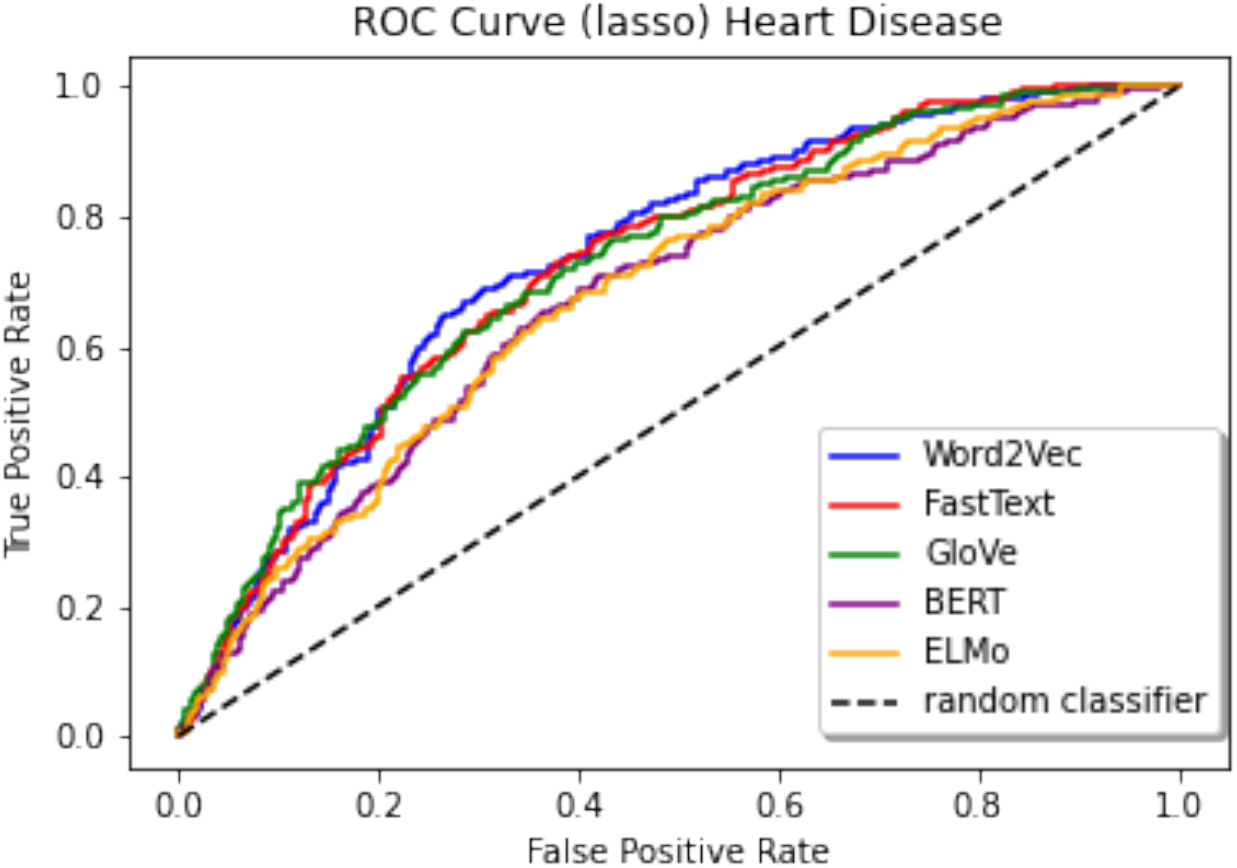
ROC curves for predicting Heart Disease (lasso)

**Fig. 14.**
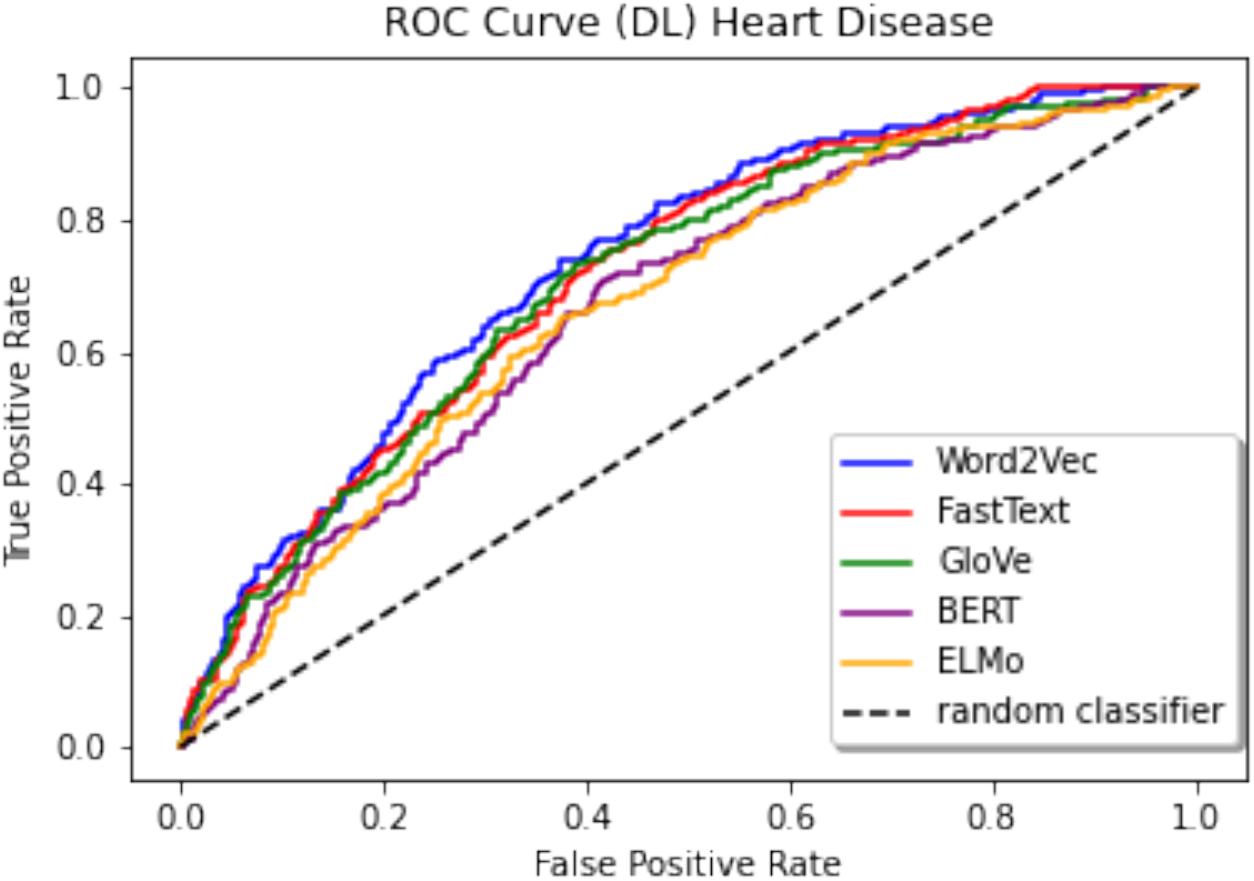
ROC curves for predicting Heart Disease (DL)

**Fig. 15.**
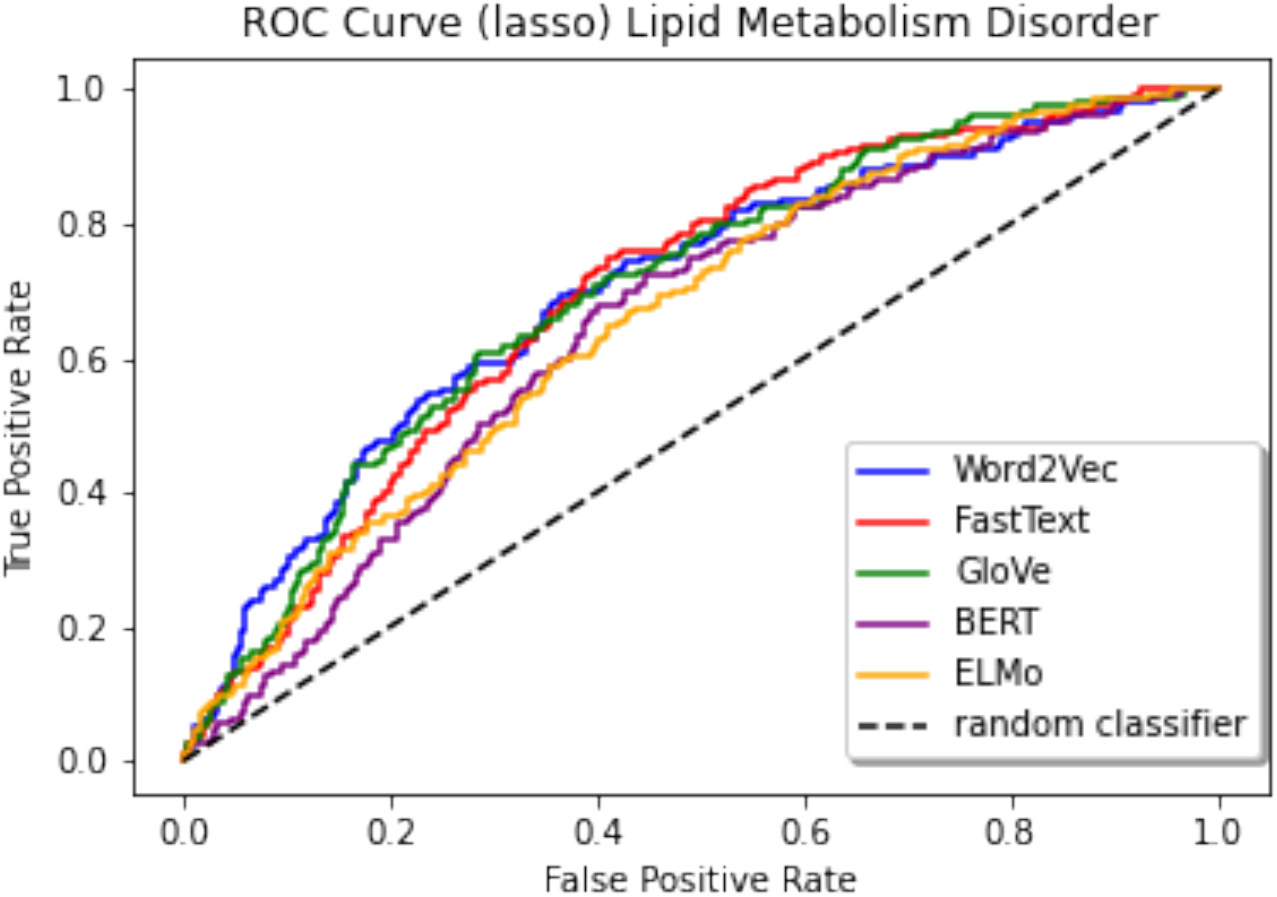
ROC curves for predicting Lipid Metabolism Disorder (lasso)

**Fig. 16.**
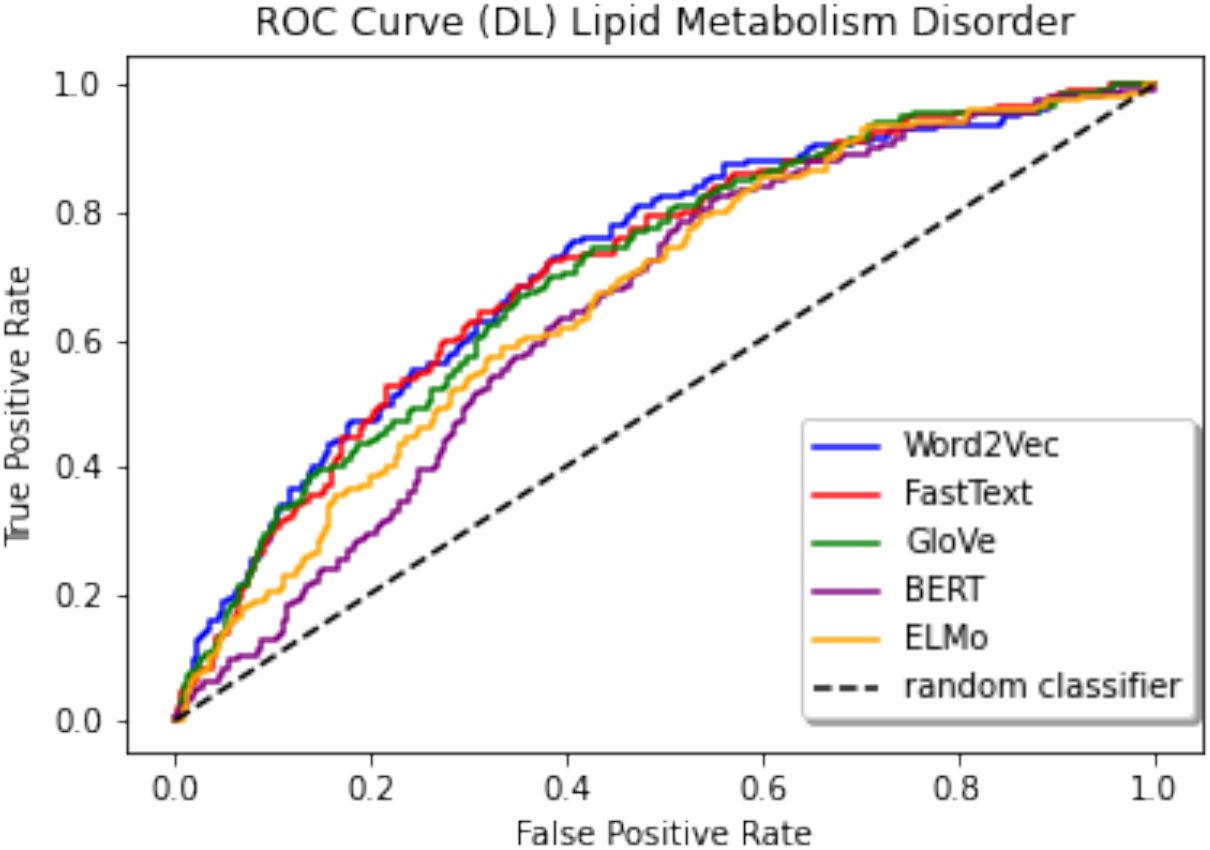
ROC curves for predicting Lipid Metabolism Disorder (DL)

Overall, it appears that the models that use Word2Vec, FastText, and GloVe embeddings tend to yield the best and most consistent results for predicting diagnoses at a patient’s next visit. The models that use ELMo static embeddings work well for some diseases, but for others, the prediction performance worsens compared to the models that use other algorithms. The models that use BERT static embeddings have the worst performance compared to models that use other algorithms. We obtain an accuracy of 0.66 for our BERT Masked Language Model, which is significantly lower than Google’s pre-trained models (ranging around 0.98). BERT is a more complex model with far more parameters than an algorithm like Word2Vec; therefore, it requires more data to build a better model. Of note, Google’s BERT models are typically trained on billions of tokens with a vocabulary (unique tokens) in tens of thousands. Our model was trained on about 1.2 million tokens with a vocabulary size of 2730. The insufficient pre-training data could be the reason for the drop in model performance. This could also be the rationale for ELMo’s worsened performance for certain diagnoses, since ELMo is a fairly complex model as well.

We also assess model performance without incorporating the temporal weights into embedding patient sequences. Table 5 shows that the time decay weights consistently improve prediction results for the diseases, but to varying degrees, noting larger improvements for predicting lipid metabolism disorder but smaller improvements for predicting cardiac dysrhythmia. Whether or not more recent events would have more of an impact on the diagnosis of a disease in the next visit would largely depend on the unique way the disease itself develops, so unsurprisingly, temporal weighting is likely more important for acute conditions than chronic conditions. Notably, many of the deep learning models have a significant drop in performance when time decay weighting is not incorporated as opposed to when it is.

**Table 5.** Prediction performance for various models that use patient vectors generated by each embedding algorithm based on static embeddings without temporal adjustment

| **Diabetes** | **Word2Vec** | **FastText** | **GloVe** | **ELMo** | **BERT** |
| --- | --- | --- | --- | --- | --- |
| AUC (lasso) | 0.6486 | 0.6395 | 0.6334 | 0.6417 | 0.6516 |
| AUC (DL) | 0.4930 | 0.4760 | 0.5203 | 0.4992 | 0.4971 |
| F1 score (lasso) | 0.2056 | 0.1731 | 0.1372 | 0.1745 | 0.0824 |
| F1 (DL) | 0.0174 | 0.0010 | 0.0627 | 0.0021 | 0.0966 |
| **Chronic Kidney Disease** |  |  |  |  |  |
| AUC (lasso) | 0.7431 | 0.7323 | 0.7197 | 0.7408 | 0.6772 |
| AUC (DL) | 0.4767 | 0.4798 | 0.4992 | 0.4935 | 0.4905 |
| F1 score (lasso) | 0.1843 | 0.1743 | 0.1973 | 0.1341 | 0.0007 |
| F1 score (DL) | 0.0000 | 0.0000 | 0.0125 | 0.0022 | 0.0000 |
| **Heart Failure** |  |  |  |  |  |
| AUC (lasso) | 0.7121 | 0.7044 | 0.7263 | 0.6790 | 0.6760 |
| AUC (DL) | 0.6530 | 0.6043 | 0.6880 | 0.4951 | 0.5018 |
| F1 score (lasso) | 0.1972 | 0.1987 | 0.2382 | 0.1980 | 0.1625 |
| F1 score (DL) | 0.2547 | 0.1063 | 0.2863 | 0.0050 | 0.0000 |
| **Lipid Metabolism Disorder** |  |  |  |  |  |
| AUC (lasso) | 0.6292 | 0.6465 | 0.6539 | 0.6065 | 0.5993 |
| AUC (DL) | 0.6027 | 0.5771 | 0.6398 | 0.5042 | 0.5009 |
| F1 score (lasso) | 0.0556 | 0.0652 | 0.0555 | 0.0444 | 0.0449 |
| F1 score (DL) | 0.1083 | 0.0520 | 0.1602 | 0.00429 | 0.0117 |
| **Fluid / Electrolyte Disorder** |  |  |  |  |  |
| AUC (lasso) | 0.6318 | 0.6071 | 0.6205 | 0.5741 | 0.6025 |
| AUC (DL) | 0.5918 | 0.5995 | 0.5894 | 0.5234 | 0.5002 |
| F1 score (lasso) | 0.3277 | 0.3277 | 0.3090 | 0.3221 | 0.3152 |
| F1 score (DL) | 0.3991 | 0.3391 | 0.41824 | 0.0692 | 0.1334 |
| **Cardiac Dysrhythmia** |  |  |  |  |  |
| AUC (lasso) | 0.6198 | 0.6102 | 0.6044 | 0.6008 | 0.5884 |
| AUC (DL) | 0.6028 | 0.5843 | 0.5901 | 0.50321 | 0.5006 |
| F1 score (lasso) | 0.0697 | 0.0748 | 0.0654 | 0.0830 | 0.0741 |
| F1 score (DL) | 0.1411 | 0.0647 | 0.2188 | 0.00521 | 0.0194 |

We assess whether or not models that use contextual embeddings generate better prediction results. Recall, some ICD9 diagnosis codes are generalized to level three specificity, which could be interpreted as having “multiple meanings”. Contextual embeddings could prove to be useful in providing some specificity to such diagnosis codes. We develop models using both BERT and ELMo static and contextual embeddings. The results in Table 6 show that using contextual embeddings tends to improve prediction results for ELMo. However, for BERT, it tends to worsen results compared to using static embeddings.

**Table 6.** Prediction performance for models that use patient vectors generated by contextual embeddings and by static embeddings

| **Diabetes** | **ELMo Cont.** | **ELMo Static** | **BERT Cont.** | **BERT Static** |
| --- | --- | --- | --- | --- |
| AUC (lasso) | 0.7235 | 0.7243 | 0.6386 | 0.6624 |
| AUC (DL) | 0.7000 | 0.6665 | 0.5963 | 0.6028 |
| F1 score (lasso) | 0.1200 | 0.0810 | 0.07921 | 0.0204 |
| F1 score (DL) | 0.2254 | 0.1542 | 0.0216 | 0.0088 |
| **Chronic Kidney Disease** |  |  |  |  |
| AUC (lasso) | 0.7590 | 0.7234 | 0.6671 | 0.6772 |
| AUC (DL) | 0.7136 | 0.7079 | 0.6365 | 0.6620 |
| F1 score (lasso) | 0.0000 | 0.0284 | 0.0425 | 0.0007 |
| F1 score (DL) | 0.2180 | 0.1089 | 0.0000 | 0.0000 |
| **Heart Failure** |  |  |  |  |
| AUC (lasso) | 0.7133 | 0.6792 | 0.6276 | 0.6739 |
| AUC (DL) | 0.6731 | 0.6594 | 0.6182 | 0.6522 |
| F1 score (lasso) | 0.1940 | 0.1404 | 0.1025 | 0.1478 |
| F1 score (DL) | 0.2670 | 0.2295 | 0.0028 | 0.0116 |
| **Lipid Metabolism Disorder** |  |  |  |  |
| AUC (lasso) | 0.6912 | 0.6600 | 0.6511 | 0.6503 |
| AUC (DL) | 0.6656 | 0.6669 | 0.6520 | 0.6447 |
| F1 score (lasso) | 0.1675 | 0.1086 | 0.0454 | 0.0442 |
| F1 score (DL) | 0.1923 | 0.1647 | 0.0122 | 0.0157 |
| **Fluid / Electrolyte Disorder** |  |  |  |  |
| AUC (lasso) | 0.6424 | 0.6194 | 0.5776 | 0.5909 |
| AUC (DL) | 0.5920 | 0.5976 | 0.5780 | 0.6003 |
| F1 score (lasso) | 0.3537 | 0.3093 | 0.2606 | 0.3083 |
| F1 score (DL) | 0.4746 | 0.4075 | 0.0851 | 0.1182 |
| **Cardiac Dysrhythmia** |  |  |  |  |
| AUC (lasso) | 0.6352 | 0.6175 | 0.5771 | 0.6077 |
| AUC (DL) | 0.5829 | 0.5887 | 0.5871 | 0.5758 |
| F1 score (lasso) | 0.0148 | 0.0568 | 0.0147 | 0.0145 |
| F1 score (DL) | 0.2773 | 0.1950 | 0.0000 | 0.0011 |

We compare our disease-specific models to the Patient Diagnosis Projection Similarity (PDPS) from [2] by comparing prediction performance for the four main diagnoses used in their work. The authors in this paper do not remove prior visits that contain the diagnosis code of interest, so we adjust our experiments to reflect this in order to directly compare with their results. The prevalence for each diagnosis is listed in Table 7. The results in Table 8 demonstrate an overall significant improvement in model performance when using disease-specific models.

**Table 7.**
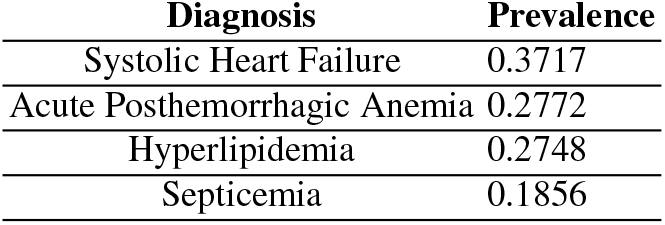
Diseases presented in the main results of [2] and their prevalences in the data

| **Diagnosis** | **Prevalence** |
| --- | --- |
| Systolic Heart Failure | 0.3717 |
| Acute Posthemorrhagic Anemia | 0.2772 |
| Hyperlipidemia | 0.2748 |
| Septicemia | 0.1856 |

**Table 8.** Comparison of PDPS prediction results from [2] to our disease-specific models. Recall that in these experiments, visits that contain instances of the target diagnosis are included in the feature window.

| **Systolic Heart Failure** | **PDPS** | **Lasso** | **DL** |
| --- | --- | --- | --- |
| AUC | 0.795 | 0.8998 | 0.9051 |
| F1 score | 0.213 | 0.7918 | 0.7994 |
| **Acute Posthemorrhagic Anemia** |  |  |  |
| AUC | 0.618 | 0.8011 | 0.7982 |
| F1 score | 0.175 | 0.7197 | 0.7035 |
| **Hyperlipidemia** |  |  |  |
| AUC | 0.723 | 0.8644 | 0.8755 |
| F1 score | 0.238 | 0.7166 | 0.7347 |
| **Septicemia** |  |  |  |
| AUC | 0.652 | 0.7971 | 0.8141 |
| F1 score | 0.242 | 0.5626 | 0.6202 |

### 3.5 Computational Efficiency

We assess computational efficiency by measuring the amount of time that it takes to train each algorithm on the MIMIC data using CPU and GPU. Our CPU cluster consists of 2 x Intel Xeon E5-2660 v3 @ 2.60 GHz CPUs with 128 GB RAM installed. And the GPU cluster contains 2 x GenuineIntel Intel(R) Xeon(R) Silver 4216 CPUs @ 2.10GHz with 192 GB RAM, as well as 4 x NVIDIA 2080 TI.

On the CPU, GloVe is the fastest model to train followed by Word2Vec, FastText, ELMo, and BERT. BERT is faster to train than ELMo on the GPU (see Table 9). It is important to consider that these are strict training times as they do not include the time it takes to set up each training before submitting to either cluster. Word2Vec, FastText, and GloVe are far easier to set up than ELMo and BERT. Static embeddings are also much faster to generate than contextual embeddings (see Table 10). For static embeddings, one only needs to extract the unique events in the dataset. On the other hand, obtaining contextual embeddings requires use of all the data since each embedding is a function of the sentence that contains it.

**Table 9.** Computing time in seconds for training each embedding algorithm using CPU and GPU

| **Embedding Algorithm** | **CPU (s)** | **GPU (s)** |
| --- | --- | --- |
| Word2Vec | 89 | 24 |
| FastText | 262 | 48 |
| GloVe | 11 | 14 |
| BERT | 1830 | 60 |
| ELMo | 382 | 72 |

**Table 10.** Computing time in seconds for generating static and contextual embeddings for ELMo and BERT using CPU and GPU

| **Type of Embedding** | **CPU (s)** | **GPU (s)** |
| --- | --- | --- |
| ELMo (contextual) | 8942 | 8688 |
| ELMo (static) | 67 | 70 |
| BERT (contextual) | 1301 | 343 |
| BERT (static) | 162 | 112 |

## 4 Discussion

In this paper, we have re-purposed traditional word embedding technologies for sequences of structured medical codes rather than actual sentences or words, and compared advanced word embedding methods for analysis of EHRs data in extensive numerical experiments. Based on our findings, for this scale of data, we would recommend Word2Vec, FastText, or GloVe to practitioners, due to their relatively fast training and set-up time, and competitive prediction performance compared to the other algorithms. ELMo and especially BERT did not perform as well in our experiments when using static embeddings, likely due to the fact that the algorithms require substantially more training data. Both of these algorithms have far more parameters than Word2Vec, GloVe, or FastText. However, the embeddings that incorporated contextual information for ELMo led to improvements in model performance for many diagnoses.

Implementing a time decay weighting did improve results to varying degrees among the diagnoses. However, the extent to which there is an improvement depends on the unique progression of a diagnosis. It could be argued that model performance for predicting other medical conditions could decrease when using the time decay weighting. For example, the development of certain cancers such as lung cancer that can be attributed to cumulative impact of risk factors such as smoking over a long period of time including particularly, early in life.

A major finding of this work is the fact that the use of contextual embeddings as opposed to static embeddings can improve model performance across many disease prediction models. We see this in our comparisons of models that utilize ELMo contextual embeddings vs. ELMo static embeddings. This displays the potential advantages of using a more advanced model that can incorporate contextual information on structured medical events. We hypothesize that given more training data, one could achieve better results than Word2Vec, GloVe, or FastText when using contextual embedding. This is likely due to the fact that algorithms such as ELMo and BERT can generate embeddings that improve the specificity of some ambiguous medical codes, thus leading to better models. However, we do not see this improvement when using BERT contextual embeddings, likely due to a lack of enough training data. BERT is a larger model than ELMo.

We found that building disease-specific models significantly outperforms the PDPS prediction method. However, both of these approaches have their own strengths and weaknesses. For a practitioner interested in predicting one disease in particular, it would be beneficial to use a disease-specific model for improved prediction accuracy. However, if one is more interested in the ability to predict risk for multiple medical conditions, PDPS would be the better method to implement due to its ability to solve a multi-classification problem. With our methods, one would need to create a new model for each new diagnosis to predict, which is more inconvenient and time-consuming.

One limitation of this work is that EHRs tend to have irregular time intervals between visits and events, but only the ordering of the events was incorporated to capture the temporal information in our experiments. As such, this approach does not necessarily paint an accurate picture of the patient record. To mitigate this problems, we could instead feed the individual event embeddings into a recurrent neural network with the exact time stamps to generate the embeddings for patient sequences before building prediction models. This way, we could more effectively utilize the temporal information and create a better model for each medical event.

There is also the issue of the time decay weighting that yields varying results depending on the diagnosis. In this paper, we use the same equation as in [2] to generate the fixed time decay weighting. It is of interest to see how varying the time decay factor could optimize prediction results for various diagnoses. This could also elucidate the unique temporal progressions of various diseases as they are compared with one another.

In this work, we are focused on comparing (unsupervised) word embedding algorithms for structured medical codes that do not require the use of a clinical outcome to train. These embeddings can be subsequently used for a wide range of downstream learning tasks such as building prediction models for multiple disease outcomes or clustering for disease subtype identification etc. The prediction models in our numerical experiments are used as an extrinsic evaluation approach [21] to compare the word embedding algorithms in the settings of our interest, rather than the focus of the paper. We recognize that there is typically a trade-off between general applicability for multiple tasks and optimal performance for a specific task. For example, there exist sophisticated prediction models for medical events that incorporate medical code embedding, for example, RETAIN, a two-level neural network attention model for sequential data [22] and GRAM: a graph-based attention model for healthcare representation learning [23]. The embeddings generated from these models are trained using a pre-specified clinical outcome through supervised learning, and may be sub-optimal for other downstream learning tasks such as predicting a different clinical outcome or multiple clinical outcomes. While it is beyond the scope of our current work, it is of future interest to compare these embeddings with those from the word-embedding methods investigated in this paper.

In addition, there are other embedding algorithms that can be used in analysis of EHRs data, e.g., Google’s Universal Sentence Encoder (USE) and Generalized Auto-Regressive model for NLU (XLNet). It is of potential interest to compare these methods in future research with more data, when it becomes possible to train these models using the MIMIC-III data.

## Data Availability

All data produced in the present study are available upon reasonable request to the authors

https://physionet.org/content/mimiciii-demo/1.4/

## Acknowledgements

This work is partly supported by NIH grant R01-GM124111. The content is the responsibility of the authors and does not necessarily represent the views of NIH.

